# Association between child marriage and intimate partner violence. A comparative analysis of Uganda Demographic Health Surveys 2011 and 2016

**DOI:** 10.1101/2023.06.19.23291619

**Authors:** Theopista Fokukora, Deda Ogum Alangea, Emefa Modey Amoah, Anthony Godi

**Affiliations:** Department of Population, Family and Reproductive Health, School of Public Health, University of Ghana, P. O. Box, LG 13, Legon, Ghana; Department of Biostatistics, School of Public Health, University of Ghana, P. O. Box, LG 13, Legon, Ghana

**Keywords:** Child Marriage, Intimate Partner Violence, Uganda Demographic and Health survey

## Abstract

The incidence of child marriage (CM) and intimate partner violence (IPV) are high in Uganda. The study sought to assess the association between child marriage and IPV across two waves of the Ugandan Demographic and Health Survey (UDHS). The evidence is important to inform policy implementation strategies aimed at curbing child marriages and IPV.

We analyzed data from UDHS 2011 and 2016. The analysis was restricted to women who have ever cohabited. Simple and multiple logistic regression models were used to assess the association between child marriage and IPV.

Following the 2011 and 2016 UDHS findings, child marriage was 54.7% in 2011 and 47.4% in 2016. Child marriage among the current adolescents increased from 9.4% in 2011 to 14.2% in 2016. According to UDHS 2011, 40.0% and 39.0% experienced physical and sexual violence respectively. In 2011 and 2016, 23.1% and 17.4% experienced both physical and sexual violence. There are higher odds of physical violence among child marriages compared to adult marriages in 2011 and 2016 (AOR = 1.44; 95% CI: 1.12-1.84 in 2011 and AOR = 1.26; 95% CI: 1.12-1.42). A wealthy or educated woman has lower odds (AOR = 0.61; 95% CI: 0.41-0.91 and AOR = 0.41; 95% CI: 0.25-0.66) to experience both physical violence and sexual violence than a poor or uneducated one. Alcohol is the most common cause of IPV. A woman married to an husband who consumes alcohol has higher odds of experiencing physical, sexual or both physical and sexual violence as compared to someone whose husband did not consume alcohol (AOR = 2.23; 95% CI:1.61-3.09 in 2011 and AOR = 2.42; 95% CI: 2.15-2.73 in 2016)

Intimate Partner Violence is associated with child marriage, level of education, wealth quintile, residence, and partner’s alcohol consumption. Much emphasis needs to be directed to cultural, as well as social economic factors but more specifically on the contribution of women emancipation/empowerment to prevent IPV.

## Introduction

Child marriage (CM) is one of the concerns of public health since below 18 years, the child’s physical, psychomotor, and cognitive developments are still developing. CM is common in both developed and developing countries. Each year globally, 15 million marry below age 18 [1, 2]. Further, studies have found out that worldwide, 5% marry below 15, and 25% marry between 15-17 years [3]. The projection is likely that up to 150 million will be married in the next decade [1]. Recent research shows that 2.2 million adolescents marry in Europe, particularly in the Central and Eastern part [4]. Child marriage is highly prevalent in sub-Saharan Africa. It is evidenced that 55% of women in Africa are married before the age of 18 [5]. The recent study carried out in Sub Saharan Africa found out that child marriage varies, it ranged from 13.5% in Rwanda to 77% in Chad [1]. A round of DHS analysis from most countries in West Africa found out that child marriage is 28% in Nigeria, 25% Burkina Faso, and 60% Niger [6]. In Ghana, 20% of girls marry by 18 years [7]. On the other hand, In Ethiopia, 40% of them were married before 18 years [8, 9].

Intimate Partner Violence (IPV) on the other hand is common in communities. Globally IPV increased from 1.1 million in 2010 to 1.4 in 2011[10]. Tia in his research found that 40% of the women who had faced IPV, only 7% had been reported [10, 11]. It is important to note that 15%-71% of the women worldwide experience a form of violence [11]. Importantly, 37% of the ever partnered partners face violence in Sub-Saharan Africa [12]. IPV is presented in different categories. Evidence has shown that physical violence accounts to 39%, psychological violence accounts to 45%, while almost a quarter (20%) face sexual violence in low income countries [13]. Systematic reviews indicate that IPV is high in Sub Saharan Africa, ranging from 43.4% in Nigeria to 76.5% in Ethiopia [14, 15]. Theoretical and empirical studies have evidenced that perpetuation of violence begins in the early childhood development [16].

Researchers have shown that CM and IPV are closely linked. Globally, 35% of young people below 18 years suffered IPV, though it’s most prevalent in East Africa 45.1% [1, 17]. One out of every four (29%) of IPV happened to those married before 18 years [4].The WHO multi-country survey about violence against women found out that IPV ranged from 19% to 66% among 15-24 years, but mostly among the adolescent marrieds than the adults [12].

Factors other than CM have been found to be associated with IPV. These include social-economic norms and beliefs, social-economic status, employment, income stress, poor communication, alcohol consumption of the husband[18], in-law violence and gender inequitable norms [19, 20].

Uganda’s CM currently has been decreasing overtime, from 56% in 2006 [38], to 49% in 2010 [39] and then 43% in 2016 [21][38]. IPV according to UDHS 2006 increased from 48% to 56% in 2011, and then reduced to 39.6% in 2016 [18, 22]. The culture in Uganda as far as IPV is concerned, is profoundly embedded in disciplining the wives so that husbands can be seen as the superiors and the controllers [23]. More than half of men and nearly a quarter of the women had attitude supportive of wife beating [21]. Recent research still confirms that IPV is social-culturally justified by both the perpetuators and the sufferer [24].

Generally, CM and IPV usually end up in complications including but not limited to injuries, depression, emotional distress often resulting in suicide [25-27]. Studies have shown that 38% of murdered women were a result of IPV [27, 28]. Sexual violence results in complications including death of child and/or mother [27], in addition to sexually transmitted diseases like HIV [29]. There is limited research on the association between CM and IPV in Uganda, which this study intends to find out. The findings of this research will be relevant for triggering policy implementation thereby improving the general health of the adolescent and thus achieving the SDG goal no 5.

## Materials and Methods

Data from the Ugandan DHS (2011 and 2016) were used in this analysis. Using a 2-stage cluster sampling, the census enumeration areas were primary sampling units while households were secondary sampling units. The 2014 population and housing census included women aged 15-49 years, permanently residing in Uganda, and these were eligible to participate. The sample weights were derived from the domestic violence sub-sample which was nationally representative. The weighted sample of 3,461 and 6,701 women was selected for the study for 2011 and 2016 respectively. The analysis included only women who had ever cohabited and had responded to the domestic violence module in survey. Women who never answered intimate partner violence questions were excluded from the study.

Ever married women and violence from an intimate partner were the variables of interest. Spousal violence was measured by modified Conflict Tactic Scale which was developed by Murray Straus in 1970s. This tool included specific questions on violence. Concerns about physical violence included; a) Push you, shake you or thrown something at you, b) Slap you, c) Twist your arm or pull your hair, d) Punch you with his fist or with something that could hurt you, e) Kick, drag, or beat you up, f)Try to choke you or burn you on purpose, g)Threatened to attack you with a knife, gun or any other weapon. A yes to one or more for the next 7 items constituted physical violence. Concerns about sexual violence included; a) Physically force you to have sexual intercourse with him even when you did not want to? b) Force you to perform any sexual acts you did not want to. A yes to one or all the two last statements constitutes sexual violence.

Age was at first cohabitation was binary grouped (below 18 years as child marriage and above 18 as adult marriage). Current married children variable was also binary grouped (below 18 years as child and above 18 as an adult). This provided a better understanding of the association between child unions and intimate partner violence. The socio-demographic variables included the level of education, wealth quintile, residence, ethnicity, religion, and alcohol intake.

### Data analysis

Data was declared as a complex survey by applying sampling weights included in the dataset. The demographic variables were analyzed using descriptive statistics to estimate the weighted percentages. The patterns of association between type of IPV, age at first cohabitation/union and other socio-demographic variables were analyzed using the complex survey design adjusted Pearson Chi-square tests. These socio-demographic variables were also analyzed against the type of IPV to determine if there were any associations. The statistical significance was set at an alpha of 0.05 (5%). The data was analyzed using Stata/SE version 16 (StataCorp, College Station, Texas, USA).

### Ethics approval and consent to participate

A written request to use the 2011 and 2016 UDHS was submitted and approved by Measure DHS. The analysis was performed in accordance with relevant regulations and guidelines. Data used in this study was anonymized before use. Since the participants in this study were below 18 years old, written consents were sought from parents/guardians while emancipated minors signed their informed consent.

## Results

The results indicated that CM was 54.7% in 2011 and 47.4% in 2016. CM among the current adolescents increased from 9.4% in 2011 to 14.2% in 2016. Most (59.4% and 57.4%) of the participants had attained primary level of education in 2011 and 2016 respectively. Majority (80.2% in 2011 and 73.3% in 2016) of the women come from rural settings. A quarter (25%) of the women belonged to the richest quartile. Uganda is predominantly a Christian country with majority of the women being Catholics and Protestants (table 1 below).

**Table 1:** Socio-demographic characteristics of women of childbearing age Uganda (UDHS 2011&2016)

|  | UDHS 2011 | UDHS 2016 |
| --- | --- | --- |
| Category | Weighted Percentage (%) | Weighted Percentage (%) |
| **Age at cohabitation** |  |  |
| Below 18 years | 54.7 | 47.4 |
| 18 years and above | 45.3 | 52.6 |
| **Current marital status among <18 years** |  |  |
| Ever married or cohabited. | 9.4 | 14.2 |
| Never married or cohabited | 90.6 | 85.8 |
| **Level of education** |  |  |
| None | 12.9 | 9.6 |
| Primary | 59.4 | 57.4 |
| Secondary | 22.5 | 25.1 |
| Higher | 5.2 | 7.9 |
| **Level of income** |  |  |
| Poorest | 17.5 | 17.5 |
| Poorer | 18.2 | 18.4 |
| Middle | 18.5 | 18.7 |
| Richer | 19.9 | 19.9 |
| Richest | 25.9 | 25.5 |
| **Place of residence** |  |  |
| Urban | 19.8 | 26.7 |
| Rural | 80.2 | 73.3 |
| **Region** |  |  |
| Kampala | 9.7 | 5.5 |
| Central 1 | 11.0 | 13.5 |
| Central 2 | 10.4 | 10.6 |
| East Central | 10.0 | 9.1 |
| Eastern | 14.6 | 17.2 |
| North | 8.5 | 10.5 |
| Karamoja | 3.3 | 2.0 |
| West-nile | 5.8 | 6.7 |
| Western | 14.1 | 12.8 |
| Southwestern | 12.7 | 12.1 |
| **Religion** |  |  |
| Catholic | 40.6 | 39.7 |
| Protestant | 30.0 | 31.2 |
| Muslim | 13.0 | 12.9 |
| Pentecostal | 13.3 | 13.3 |
| SDA | 1.9 | 1.7 |
| Others | 1.2 | 12.4 |
| **TOTAL** | **100** | **100** |

Bivariate analysis (in table 2 below) indicated that 40.3% in 2011 and 39.0% in 2016 of the women experienced physical violence. A quarter (27.5% in 2011 and 22.5% in 2016) of the women experienced sexual violence, while 19.2% 16.0% of the women experienced both physical and sexual violence in 2011 and in 2016 respectively. Child marriage was significantly associated with IPV (p<0.05). Physical violence among child marriages was higher as compared with adult marriages (45.4% vs 34.2% in 2011 and 44.2% vs 34.3% in 2016; p<0.05). Similarly, child marriage experienced a both physical and sexual violence as compared to adult marriages (23.1% vs 14.4% in 2011 and 17.4% vs 14.7% in 2016; p<0.01).

**Table 2:** Bivariate analysis of the experience of IPV across different socio-demographic groups

|  |  |  | UDHS 2011 2011 |  |  | UDHS 2016 |  |  |
| --- | --- | --- | --- | --- | --- | --- | --- | --- |
| Variables |  |  | Physical violence<br>% (x<sup>2</sup>) | Sexual violence<br>% (x<sup>2</sup>) | Physical & Sexual<br>Violence % (x<sup>2</sup>) | Physical violence<br>% (x<sup>2</sup>) | Sexual violence<br>% (x<sup>2</sup>) | Physical and Sexual<br>violence % (x<sup>2</sup>) |
| **Age at cohabitation (years)** | 18 and above |  | 34.2 (22.27) *** | 22.9 (15.31) * | 14.4 (20.56) *** | 34.3 (77.89) *** | 22.2 (0.60) | 14.7 (9.67) ** |
|  | Below 18 |  | 45.4 | 31.3 | 23.1 | 44.2 | 22.9 | 17.4 |
| **Current age (years)** | 18 and above |  | 40.8 (6.99) | 27.6 (2.26) | 19.3 (0.92) | 39.3 (11.07) ** | 22.7 (6.12) * | 16.1 (6.82) * |
|  | Below 18 |  | 19.1 | 17.0 | 13.0 | 25.1 | 13.6 | 7.8 |
| **Level of education** | None |  | 45.8 (49.09) * | 22.1 (21.52) * | 19.1 (17.69) ** | 47.2 (221.56) *** | 21.3 (73.38) *** | 17.8 (66.70) ** |
|  | Primary |  | 44.2 | 31.1 | 21.8 | 43.1 | 25.1 | 18.0 |
|  | Secondary |  | 29.3 | 23.5 | 13.0 | 29.5 | 20.0 | 12.4 |
|  | Post-secondary |  | 13.8 | 14.9 | 8.9 | 17.5 | 9.8 | 6.4 |
| **Level of income** | Poorest |  | 53.5 (80.58) * | 31.4 (20.11) * | 27.4 (32.16) *** | 50.2 (217.94) *** | 23.2(59.58) *** | 18.8 (53.01) *** |
|  | Poorer |  | 48.5 | 28.5 | 21.8 | 43.8 | 25.9 | 18.8 |
|  | Middle |  | 41.5 | 31.0 | 20.1 | 39.6 | 24.6 | 15.8 |
|  | Richer |  | 35.2 | 25.6 | 15.8 | 36.2 | 23.9 | 16.3 |
|  | Richest |  | 24.5 | 22.0 | 11.7 | 25.8 | 15.6 | 10.6 |
| **Place of residence** | Urban |  | 29.9 (15.61) ** | 24.2 (2.00) | 13.2 (7.61) * | 32.0 (49.31) *** | 18.9 (18.56) ** | 13.8 (8.79) ** |
|  | Rural |  | 42.4 | 28.2 | 20.3 | 41.2 | 23.7 | 16.7 |
| **Region** | Kampala |  | 28.8 (87.63) * | 21.5 (45.93) | 10.9 (27.07) * | 24.8 (169.62) *** | 14.4 (264.24) *** | 10.1 (155.65) *** |
|  | Central |  | 28.4 | 25.1 | 15.7 | 32.7 | 19.4 | 14.4 |
|  | Central |  | 30.6 | 33.6 | 16.4 | 31.0 | 18.0 | 11.3 |
|  | East Central |  | 35.1 | 32.8 | 18.1 | 31.3 | 24.6 | 14.8 |
|  | Eastern |  | 57.2 | 34.4 | 28.8 | 49.3 | 44.8 | 16.1 |
|  | North |  | 55.3 | 27.8 | 22.3 | 41.7 | 22.6 | 18.1 |
|  | Karamoja |  | 36.0 | 16.4 | 15.6 | 41.1 | 16.9 | 11.9 |
|  | West-Nile |  | 46.3 | 27.7 | 23.9 | 45.4 | 21.7 | 17.5 |
|  | Western |  | 40.5 | 25.4 | 18.1 | 37.5 | 30.3 | 18.8 |
|  | Southwestern |  | 37.9 | 20.6 | 16.9 | 43.2 | 26.1 | 19.5 |
| **Religion** | Catholic |  | 44.2 (17.35) | 27.4 (8.54) | 21.1 (8.02) | 43.5 (70.72) * | 22.1 (5.03) | 16.6 (11.40) |
|  | Protestant |  | 36.6 | 25.9 | 16.6 | 38.7 | 23.4 | 16.7 |
|  | Muslim |  | 33.4 | 31.7 | 18.1 | 31.1 | 21.6 | 13.4 |
|  | Pentecostal |  | 44.7 | 28.4 | 20.3 | 36.4 | 23.5 | 16.1 |
|  | SDA |  | 35.7 | 13.9 | 10.6 | 22.0 | 20.1 | 8.8 |
|  | Others |  | 41.1 | 32.8 | 32.8 | 31.2 | 16.0 | 13.0 |
| **Husband' takes Alcohol** | No |  | 30.3 (301.53) *** | 24.7 (214.55) *** | 14.0 (33.73) *** | 28.2 (484.37) *** | 17.9 (119.95) *** | 10.8 (198.70) *** |
|  | Yes |  | 51.6 | 30.8 | 25.1 | 53.2 | 28.6 | 22.8 |
| **Overall Percentage score** |  |  | **40.3** | **27.5** | **19.2** | **39.0** | **22.5** | **16.0** |

\*, \*\*, \*\*\* corresponds with p<0.05, p<0.01, and p<0.001 respectively

For the section of currently married teenagers (below 18 years), a substantial proportion (19.1% in 2011 and 25.1% in 2016, p<0.001) suffer physical violence. Exactly 17.0% in 2011 and 13.6% in 2016 suffer sexual violence (p<0.05). The proportion of those among this group who suffered both physical and sexual violence reduced from 13% in 2011 to 7.8% in 2016 (p<0.05).

Almost half of women who experienced physical violence had no formal education (45.8% in 2011 to 47.2% in 2016). Experience of physical violence reduced with higher levels of education (p<0.05). Rural women experience twice as much physical violence more than the urban settings (p<0.001). Women from Eastern, North and Karamoja region experience more physical violence compared to those form other regions (p<0.001). While the physical violence reduced among women with non-alcoholic husbands (30.3% in 2011 to 28.3% in 2016), it increased among those whose spouses drunk alcohol (51.6% in 2011 to 53.2% in 2016; p<0.001).

The results also indicated that sexual violence reduced among other levels of education except in no formal education category (p<0.01). The higher wealth quintile was observed to reduce sexual violence among the women (p<0.01). Rural women experience more sexual violence compared to women urban settings (p<0.01). Sexual violence increased among women from Eastern, North and Karamoja region compared to those form other regions (p<0.001). Whereas the sexual violence greatly reduced among women with non-alcoholic spouses (24.7% in 2011 to 17.9% in 2016 p<0.001), it slightly reduced among those with alcoholic husbands (30.8% in 2011 to 28.6% in 2016; p<0.001).

Taking both physical and sexual violence into account, the results indicated that higher wealth and education levels were associated with the lower risk of the combined type of violence (p<0.01). Furthermore, both physical and sexual violence more than doubled among the women whose husbands take alcohol as compared to those whose husbands were not alcoholics (25.1% vs 14.0% in 2011 and 22.8% vs 10.8% in 2016; p<0.0001).

CM recorded significantly higher odds of physical violence as compared to adult marriages in both 2011 (AOR = 1.44; 95% CI: 1.12-1.84) and 2016 (AOR = 1.26; 95% CI: 1.12-1.42). The odds of experiencing physical violence reduces with increasing level of education and wealth index. An educated woman had lower odds of experiencing physical violence as compared to uneducated woman in 2016 as compared to 2011 (AOR = 0.42; 95% CI: 0.17-1.06 in 2011 to AOR = 0.37; 95% CI: 0.27-0.52; p<0.05 in 2016).

Conversely, the odds of experiencing physical violence among wealthy women were lower compared to a poor woman, however, there was an increased in IPV among wealthy women in 2016 when compared to 2011 (AOR = 0.46; 95% CI: 0.25-0.83 in 2011 to AOR = 0.61; 95% CI: 0.47-0.80; p<0.05 in 2016). The women in the Eastern regions had higher odds of experiencing both physical and sexual violence as compared to other regions (AOR=2.85; 95% CI:1.70-4.76). Women whose husbands drink alcohol had higher odds of experiencing both physical and sexual violence as compared to those of a woman whose husbands were non-alcoholics (AOR = 2.23; 95% CI:1.61-3.09 in 2011 and AOR = 2.42; 95% CI: 2.15-2.73 in 2016) (table 3).

**Table 3:**
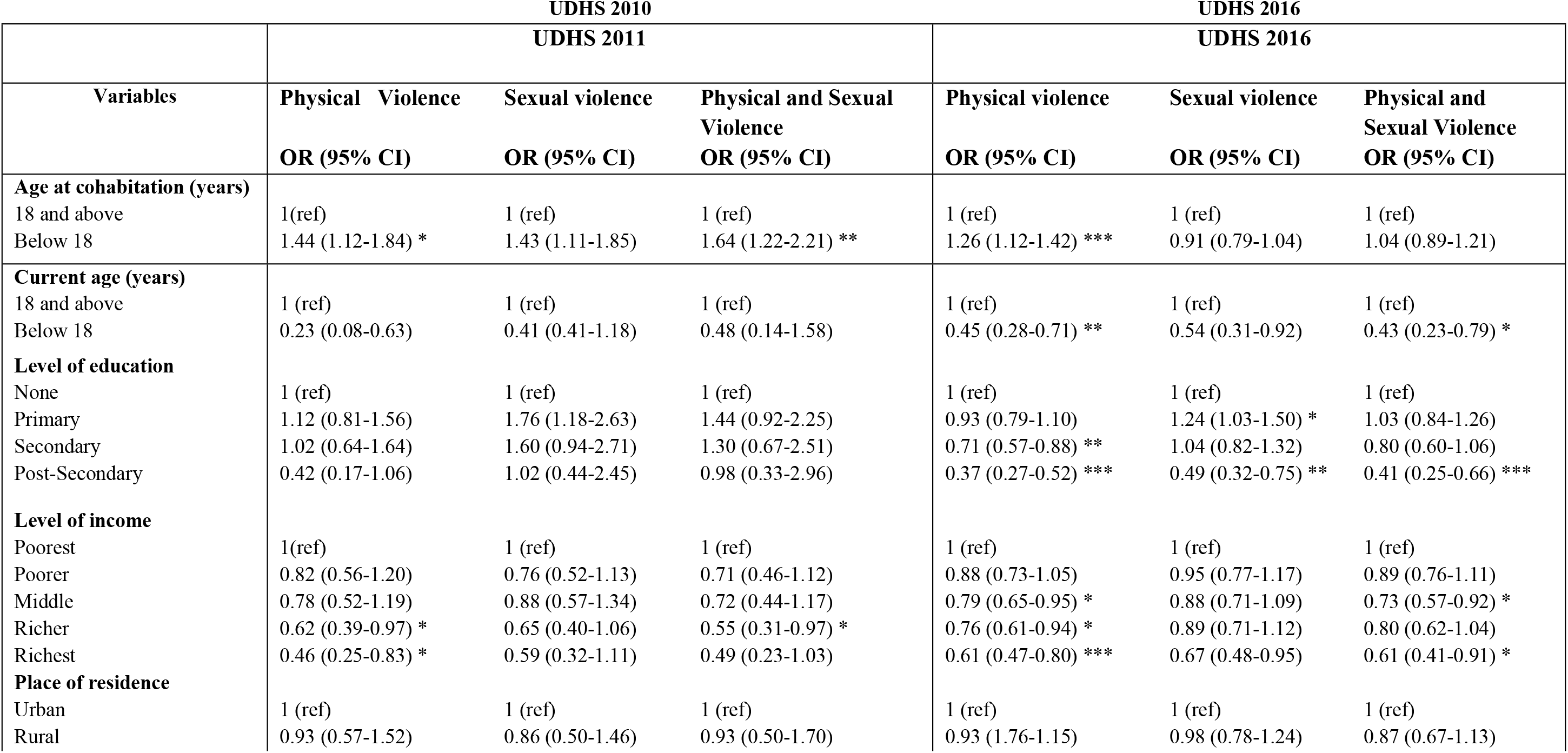
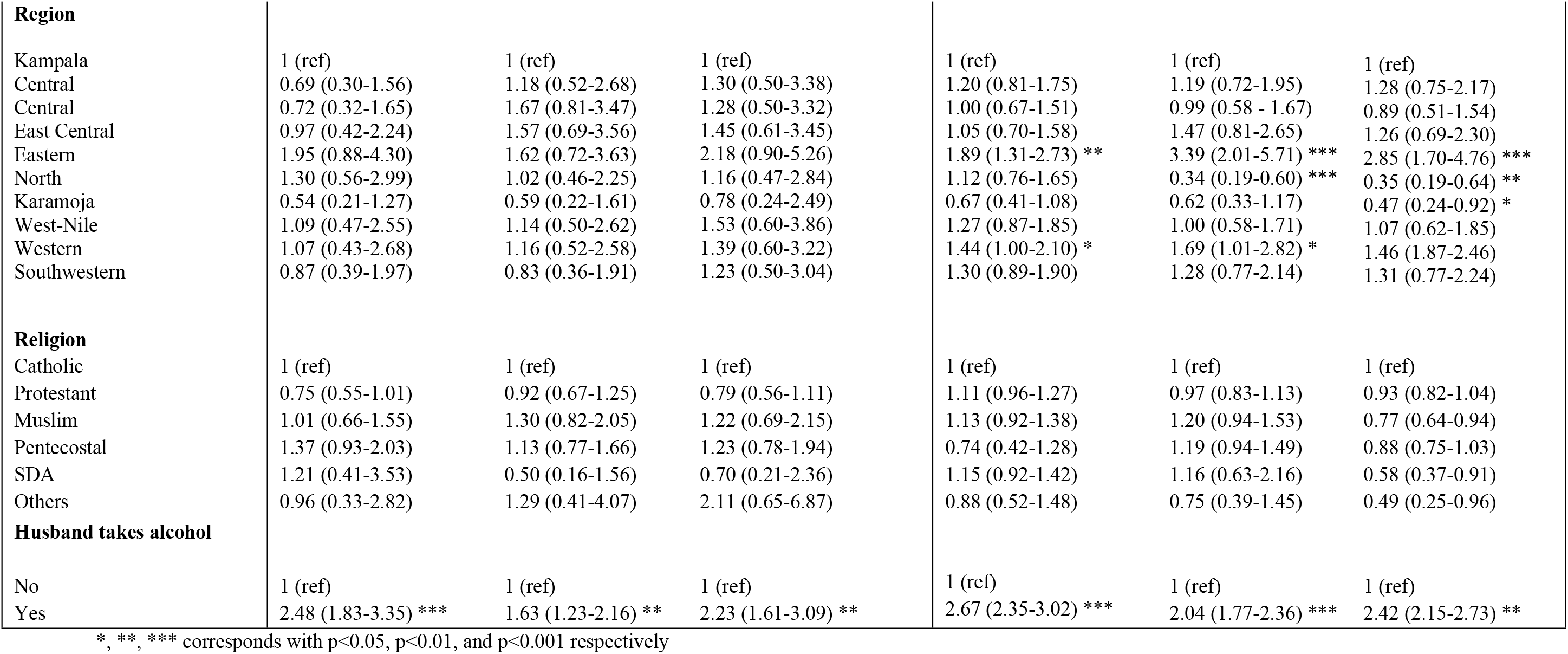
Logistic regression models showing adjusted OR between demographic characteristics and Different types of violence in Uganda (UDHS 2011 & 2016)

|  | UDHS 2010<br>UDHS 2011 |  |  | UDHS 2016<br>UDHS 2016 |  |  |
| --- | --- | --- | --- | --- | --- | --- |
| Variables | Physical Violence<br>OR (95% CI) | Sexual violence<br>OR (95% CI) | Physical and Sexual Violence<br>OR (95% CI) | Physical violence<br>OR (95% CI) | Sexual violence<br>OR (95% CI) | Physical and Sexual Violence<br>OR (95% CI) |
| **Age at cohabitation (years)** |  |  |  |  |  |  |
| 18 and above | 1 (ref) | 1 (ref) | 1 (ref) | 1 (ref) | 1 (ref) | 1 (ref) |
| Below 18 | 1.44 (1.12-1.84) * | 1.43 (1.11-1.85) | 1.64 (1.22-2.21) ** | 1.26 (1.12-1.42) *** | 0.91 (0.79-1.04) | 1.04 (0.89-1.21) |
| **Current age (years)** |  |  |  |  |  |  |
| 18 and above | 1 (ref) | 1 (ref) | 1 (ref) | 1 (ref) | 1 (ref) | 1 (ref) |
| Below 18 | 0.23 (0.08-0.63) | 0.41 (0.41-1.18) | 0.48 (0.14-1.58) | 0.45 (0.28-0.71) ** | 0.54 (0.31-0.92) | 0.43 (0.23-0.79) * |
| **Level of education** |  |  |  |  |  |  |
| None | 1 (ref) | 1 (ref) | 1 (ref) | 1 (ref) | 1 (ref) | 1 (ref) |
| Primary | 1.12 (0.81-1.56) | 1.76 (1.18-2.63) | 1.44 (0.92-2.25) | 0.93 (0.79-1.10) | 1.24 (1.03-1.50) * | 1.03 (0.84-1.26) |
| Secondary | 1.02 (0.64-1.64) | 1.60 (0.94-2.71) | 1.30 (0.67-2.51) | 0.71 (0.57-0.88) ** | 1.04 (0.82-1.32) | 0.80 (0.60-1.06) |
| Post-Secondary | 0.42 (0.17-1.06) | 1.02 (0.44-2.45) | 0.98 (0.33-2.96) | 0.37 (0.27-0.52) *** | 0.49 (0.32-0.75) ** | 0.41 (0.25-0.66) *** |
| **Level of income** |  |  |  |  |  |  |
| Poorest | 1 (ref) | 1 (ref) | 1 (ref) | 1 (ref) | 1 (ref) | 1 (ref) |
| Poorer | 0.82 (0.56-1.20) | 0.76 (0.52-1.13) | 0.71 (0.46-1.12) | 0.88 (0.73-1.05) | 0.95 (0.77-1.17) | 0.89 (0.76-1.11) |
| Middle | 0.78 (0.52-1.19) | 0.88 (0.57-1.34) | 0.72 (0.44-1.17) | 0.79 (0.65-0.95) * | 0.88 (0.71-1.09) | 0.73 (0.57-0.92) * |
| Richer | 0.62 (0.39-0.97) * | 0.65 (0.40-1.06) | 0.55 (0.31-0.97) * | 0.76 (0.61-0.94) * | 0.89 (0.71-1.12) | 0.80 (0.62-1.04) |
| Richest | 0.46 (0.25-0.83) * | 0.59 (0.32-1.11) | 0.49 (0.23-1.03) | 0.61 (0.47-0.80) *** | 0.67 (0.48-0.95) | 0.61 (0.41-0.91) * |
| **Place of residence** |  |  |  |  |  |  |
| Urban | 1 (ref) | 1 (ref) | 1 (ref) | 1 (ref) | 1 (ref) | 1 (ref) |
| Rural | 0.93 (0.57-1.52) | 0.86 (0.50-1.46) | 0.93 (0.50-1.70) | 0.93 (1.76-1.15) | 0.98 (0.78-1.24) | 0.87 (0.67-1.13) |

| **Region** |  |  |  |  |  |  |
| --- | --- | --- | --- | --- | --- | --- |
| Kampala | 1 (ref) | 1 (ref) | 1 (ref) | 1 (ref) | 1 (ref) | 1 (ref) |
| Central | 0.69 (0.30-1.56) | 1.18 (0.52-2.68) | 1.30 (0.50-3.38) | 1.20 (0.81-1.75) | 1.19 (0.72-1.95) | 1.28 (0.75-2.17) |
| Central | 0.72 (0.32-1.65) | 1.67 (0.81-3.47) | 1.28 (0.50-3.32) | 1.00 (0.67-1.51) | 0.99 (0.58 - 1.67) | 0.89 (0.51-1.54) |
| East Central | 0.97 (0.42-2.24) | 1.57 (0.69-3.56) | 1.45 (0.61-3.45) | 1.05 (0.70-1.58) | 1.47 (0.81-2.65) | 1.26 (0.69-2.30) |
| Eastern | 1.95 (0.88-4.30) | 1.62 (0.72-3.63) | 2.18 (0.90-5.26) | 1.89 (1.31-2.73) ** | 3.39 (2.01-5.71) *** | 2.85 (1.70-4.76) *** |
| North | 1.30 (0.56-2.99) | 1.02 (0.46-2.25) | 1.16 (0.47-2.84) | 1.12 (0.76-1.65) | 0.34 (0.19-0.60) *** | 0.35 (0.19-0.64) ** |
| Karamoja | 0.54 (0.21-1.27) | 0.59 (0.22-1.61) | 0.78 (0.24-2.49) | 0.67 (0.41-1.08) | 0.62 (0.33-1.17) | 0.47 (0.24-0.92) * |
| West-Nile | 1.09 (0.47-2.55) | 1.14 (0.50-2.62) | 1.53 (0.60-3.86) | 1.27 (0.87-1.85) | 1.00 (0.58-1.71) | 1.07 (0.62-1.85) |
| Western | 1.07 (0.43-2.68) | 1.16 (0.52-2.58) | 1.39 (0.60-3.22) | 1.44 (1.00-2.10) * | 1.69 (1.01-2.82) * | 1.46 (1.87-2.46) |
| Southwestern | 0.87 (0.39-1.97) | 0.83 (0.36-1.91) | 1.23 (0.50-3.04) | 1.30 (0.89-1.90) | 1.28 (0.77-2.14) | 1.31 (0.77-2.24) |
| **Religion** |  |  |  |  |  |  |
| Catholic | 1 (ref) | 1 (ref) | 1 (ref) | 1 (ref) | 1 (ref) | 1 (ref) |
| Protestant | 0.75 (0.55-1.01) | 0.92 (0.67-1.25) | 0.79 (0.56-1.11) | 1.11 (0.96-1.27) | 0.97 (0.83-1.13) | 0.93 (0.82-1.04) |
| Muslim | 1.01 (0.66-1.55) | 1.30 (0.82-2.05) | 1.22 (0.69-2.15) | 1.13 (0.92-1.38) | 1.20 (0.94-1.53) | 0.77 (0.64-0.94) |
| Pentecostal | 1.37 (0.93-2.03) | 1.13 (0.77-1.66) | 1.23 (0.78-1.94) | 0.74 (0.42-1.28) | 1.19 (0.94-1.49) | 0.88 (0.75-1.03) |
| SDA | 1.21 (0.41-3.53) | 0.50 (0.16-1.56) | 0.70 (0.21-2.36) | 1.15 (0.92-1.42) | 1.16 (0.63-2.16) | 0.58 (0.37-0.91) |
| Others | 0.96 (0.33-2.82) | 1.29 (0.41-4.07) | 2.11 (0.65-6.87) | 0.88 (0.52-1.48) | 0.75 (0.39-1.45) | 0.49 (0.25-0.96) |
| **Husband takes alcohol** |  |  |  |  |  |  |
| No | 1 (ref) | 1 (ref) | 1 (ref) | 1 (ref) | 1 (ref) | 1 (ref) |
| Yes | 2.48 (1.83-3.35) *** | 1.63 (1.23-2.16) ** | 2.23 (1.61-3.09) ** | 2.67 (2.35-3.02) *** | 2.04 (1.77-2.36) *** | 2.42 (2.15-2.73) ** |

\*, \*\*, \*\*\* corresponds with $p < 0.05$ , $p < 0.01$ , and $p < 0.001$ respectively

## Discussion

The findings from this analysis indicated that CM reduced from 54.7% in 2011 to 47.4% in 2016. It is true and evident that CM still exist in Uganda. The Uganda Demographic and Health Survey (UDHS) findings indicate that CM currently has been decreasing overtime, from 56% in 2006 [22]. Uganda also contributes to the overall global findings whereby, 15 million marry below age 18 [1, 3]. As reflected, Sub-Saharan Africa has the highest proportion [3, 5, 30]. Evidence is derived from UNFPA and UNICEF database where, East and Southern Africa had 36% of child marriage [1]. The rate of child marriage in Uganda’s is much higher than that of Ghana, where 1 in every 5 girls marry by 18 years [10]. Recent research reveals that child marriage in Rwanda was lower (13.5%), while Chad’s proportion was high at 77% compared to Uganda’s situation [31].

The findings from this study further indicated that both physical and sexual violence reduced among child marriages from 23.1% in 2011 to 17.4% in 2016. This could have been possible because of human rights implementation programs about adolescents. Elsewhere, similar results revealed that IPV ranges from 15% -71% in different countries, but most prevalent among CM [10, 11]. In agreement with the results from this study, different kinds of violence include physical violence, psychological/emotional violence, and sexual violence accounting 39%, 45% and 20% respectively [13, 17], of which married children are most vulnerable. There is a high prevalence (76.5% and 43.4%) of IPV in Ethiopia and Nigeria respectively [18–20], which is proportional to child marriage in those countries. Recent estimates show that IPV ranged from 17.5% in Mozambique to 42% in Uganda [31]. Though such combined violence reduced among the CM, more strategies targeting communities still need to be employed to eliminate such suffering among this vulnerable population.

This study found that the odds of experiencing IPV among married children was higher as compared to married adults. The communities in Uganda discipline children because of the belief that they are not yet mature to make their own decisions. In this respect, husbands to these children use threats and beatings to justify their dominance over them. The results agree with a multi-country analysis of SSA DHS data [31]. Peterman and his colleagues (2015) while identifying the global trends, found that 1/3 of the women who experienced physical or sexual abuse were initiated to sex at a teenage age [15]. At such a tender age, the adolescents do not reveal their grievances because they can’t fight for their rights. Evidence has been found in a comparative study of 34 countries, where 29% of IPV happened to those married before 18, but 15-17 years were more at risk [3]. The results are in agreement with systematic reviews carried out in most low and middle income countries, revealing that 19% to 66% among 15-24 years were at a high risk of IPV as compared to adults [12]. Rural communities especially in Uganda need to be sensitized about helping such children to mature into responsible adults without necessarily inflicting pain to them.

The findings indicated that the level of education was associated to IPV. The higher level of education exposes the women to understanding their rights and gives them empowerment. It is likely that the individual who goes through higher levels of education, and attains employment, sustains her economic income, which reduces violence in the households. This is not the case with married children. DHS 2016 shows that women who had a low level of education were more prone to IPV [26, 32]. The results further agree with findings from a meta-analysis in Sub-Saharan Africa, which identified that the women who were not educated were more likely to face IPV [33]. Jesmin (YEAR??) identified however, that lower education was not necessarily associated with IPV [34].

Apart from education, but closely related, the level of income plays a vital role in women’s risk for IPV. Our findings indicated that the odds of IPV reduced among wealthier as compared to poor women in both 2011 and 2016. The married children are dependent on their husbands, and because of the economic breakdown especially among the rural men, this induces wife beating and other forms of violence to them. This is supported by evidence from Kenya, Zimbabwe, Nigeria [14, 35, 36], and Malawi [37]. However, other research have found that high social economic class and women empowerment, if not well managed, results in misunderstandings in the couples ending up in IPV [19, 20].

Apart from the socio-economic situation, the study indicated that residence was significantly associated with IPV and CM. Most of the children who were married were predominantly from rural settings where culturally wife beating is justified as a disciplinary behaviour. Findings indicated that rural women experienced twice as much physical violence than those in urban settings. In villages and humanitarian settings especially Somalia and South Sudan which still adore and embrace culture, IPV among such rural communities was most prevalent there [38]. This could also be related to religiosity’s fear to divorce as they would be seen as failures in churches and communities. Evidence from diverse researchers found that individual, relationship, community and societal factors were all associated with IPV [13, 39]. A recent study in Ghana identified that IPV is justified by beating, dominance, and social cultural justification [40]. The drivers to IPV were poverty, low education levels, and limited implementation of policy frameworks to promote human rights [21, 41]. These are similar scenarios identified in Uganda.

Partner alcohol intake was significantly associated with IPV in this study. This finding supports earlier findings in Uganda. A drunkard is believed to have less insight of the outcome of violence, and, men drink to supplement their status of superiority, manhood and dominance [18, 42]. The results agree with findings in Nepal - that alcohol consumption by the husband predisposed a partner to IPV [43]. The consistent drinking of alcohol is often associated with poverty. Alcohol intake further predisposes to sexual violence and uncontrolled birth rates resulting in high fertility rate.

Some limitations were identified in this study. The data was retrospective in nature and thus subject to some level of recall bias. Further, the women who report violence are often socially excluded for exposing the family affairs to the public. This could have introduced the desirability bias because of its sensitivity. Further, data used for this study was quantitatively collected, therefore some deeper expressions and experiences of women who face violence could have been missed. The qualitative approach could also reveal household and community factors.

## Conclusion and implication

The study found out that child marriage and IPV are prevalent in Uganda. There was a significant association between CM and IPV. Married children are characterized by low level of education, poor wealth quintile, and dependability. All these were significantly associated with IPV. Because IPV among adolescents has detrimental complications, policy makers and implementors need to put into consideration children rights to protect those who get married from facing this violence in developing countries like Uganda. The implementation of these rights needs to begin from the grassroot level (communities) up to the national and international agencies with an aim to improve children’s health and strengthen the awareness of children’s rights in the communities.

## Data Availability

at http://www.dhsprogram.com/data/dataset

## Acknowledgments

I acknowledge the support by the HRP Alliance, part of the UNDP-UNFPA-WHO-World Bank Special programme of Research Development and Research training in Human Reproduction (HRP), a co-sponsored programme executed by the World Health Organization (WHO) who facilitated my PhD studies. The University of Ghana, School of Public Health for their academic support in the manuscript development and submission process.

## Authors’ contributions

**Conceptualization:** Theopista Fokukora, Deda Alangea, Emefa Modey Amoah

**Formal analysis:** Theopista Fokukora, Anthony Godi

**Methodology:** Theopista Fokukora

**Supervision:** Deda Alangea, Emefa Modey Amoah, Anthony Godi

**Writing – original draft:** Theopista Fokukora

**Writing – review & editing:** Deda Alangea, Emefa Moday, Anthony Godi

## Declarations

### Consent for publication

Not applicable

### Availability of data and materials

The data is available on request at http://www.dhsprogram.com/data/dataset. The authors confirm that they followed general rules of requesting for the data from the above website, therefore did not have any special access or privileges to the data.

## Abbreviations

CM: Child marriage
IPV: Intimate Partner Violence
OR: Odds Ratio
UDHS: Uganda Demographic and Health Survey
WHO: World Health Organization

